# The State of Ohio Adversity and Resilience (SOAR) Study Protocol: A Comprehensive, Multimodal, Family-based, Longitudinal Investigation of Risk and Resilience in Mental Health and Substance Use Disorders

**DOI:** 10.1101/2024.11.19.24316679

**Authors:** Anthony P. King, Scott A. Langenecker, Stephanie Gorka, Jessica Turner, Lei Wang, Heather Wastler, Marybel R Gonzalez, Canada Keck, Randall Olsen, Hyoshin Kim, Brett Klamer, Cal Adler, Elissar Andari, Stacey L. Barrenger, Natalie Bonfine, Melanie Bozzay, Susan L. Brown, Chris Browning, Katie Burkhouse, Kathleen Carter, Kim M. Cecil, Karin Coifman, Timothy N. Crawford, Cory E. Cronin, Melissa DelBello, Steven W. Evans, Chris Flask, K. Jean Forney, Larrilyn Grant, John Gunstad, Paul J. Hershberger, Kristen R. Hoskinson, Christina Klein, Jose Moreno, Molly McVoy, Paula K. Miller, Eric E. Nelson, Randy Nesse, Chris Nguyen, Kei Nomaguchi, Alissa Paolella, Edison Perdomo, Ruchika Prakash, Colin Odden, Martha Sajatovic, Robert Smith, Jonathan Trauth, Ivy Tso, Xin Wang, Jennifer T. Grant Weinandy, Krista K. Westrick-Payne, Greta Winbush, Brian Wymbs, Hong Xie, Stephen Gavazzi, Timothy R Huerta, Grace Wentzel, Nina V. Kraguljac, K. Luan Phan

## Abstract

**Introduction:** Deaths related to drug overdose and suicide in the United States have increased 500% and 35%, respectively, over the last two decades. The human and economic costs to society associated with these “deaths of despair” are immense. Great efforts and substantial investments have been made in treatment and prevention, yet these efforts have not abated these increasing trajectories of deaths over time.The COVID-19 pandemic has exacerbated and highlighted these problems. Notably, some geographical areas (e.g. Appalachia, farmland) and some communities (e.g. low-income persons, “essential workers”, minoritized populations) have been disproportionately affected. Risk factors have been identified for substance use and suicide deaths: forms of adversity, neglect, opportunity indexes, and trauma. Yet, the biological, psychological, and social mechanisms driving risk are not uniform. Notably, most people exposed to risk factors do not become symptomatic and could broadly be considered resilient. Achieving a better understanding of biological, psychological, and social mechanisms underlying both pathology and resilience will be crucial for improving approaches for prevention and treatment and creating precision medicine approaches for more efficient and effective treatment.

**Methods and analysis:** The State of Ohio Adversity and Resilience (SOAR) study is a prospective, longitudinal, multimodal, integrated familial study designed to identify biological, psychological, and social risk and resilience factors and processes leading to disorders of the brain, including overdose, suicide and psychological/medical comorbidity (e.g., alcoholism) that are associated with alcohol use disorder whichdreducedreducedreduced life expectancy and quality of life. It includes two nested longitudinal samples: (i) Wellness Discovery Survey: an address-based random population epidemiological sample of 15,000 individuals (unique households) representative of the state of Ohio assessed for psychosocial, psychiatric, and substance use factors, and (ii) Brain Health Study: a family-based, multimodal, deep-phenotyping study conducted in 1200 families (up to 3600 persons aged 12-72) including MRI, EEG, blood biomarkers, psychiatric diagnostic interviews, as well as neuropsychological, psychosocial functioning, and family / community history, dynamics, and support assessments. SOAR is designed to discover, develop and deploy advanced predictive analytics and interventions to transform mental health prevention, diagnosis, treatment, and recovery.

**Ethics and dissemination:** All participants will provide written informed consent (or assent for minors). The study was approved by The Ohio State University Institutional Review Board (study numbers 2023H0316 and 2023H0350). Findings will be disseminated to academic peers, clinicians and healthcare consumers, policymakers, and the general public, using local and international academic channels (academic journals, evidence briefs and conferences) and outreach (workshops and seminars).

**STRENGTHS AND LIMITATIONS OF THIS STUDY:**

- This study is a unique combination of a large address-based (ABS) epidemiological random sample Wellness Discovery survey (N=15,000) representative of the State of Ohio, and a nested multimodal deep neurophenotyping study (N=3,600).
- The Brain Health neurophenotyping study is nested in families, allowing for direct study of family dynamics, contextual factors, and intergenerational transmission and redirection from mental illness.
- The neurophenotyping study includes advanced multimodal MRI and EEG at five fixed sites (and one rotating mobile site) with centralized data management and advanced standardized acquisition, multisite harmonization, pre-processing, and analyses pipelines.
- We intend cross-saturation of the Wellness Discovery Survey and Brain Health samples over time. At present, overlap is minimal due to time and funding limitations.
- We have an ambitious plan to actively engage communities across the state in two-way communication over time in this study. Due to time limitationslimitation, the first wave of data collection did not involve community-based participatory research.

## INTRODUCTION

Deaths related to drug overdoses and suicides have risen dramatically in recent decades across the USA (National Center for Health Statistics, 2024, Friedman et al., 2023, Fujita-Imazu et al., 2023). While there has been a decrease (∼3%) in the number of overdose deaths in 2023, a very high number of these deaths continue (estimated at 107,543 overdose deaths in 2023), despite substantial expenditures and tractable and creative efforts in mental health care, intervention, and prevention (Rockett et al., 2023, Lou et al., 2021). The state of Ohio also has documented substantial increases in deaths related to overdose and suicide (Health Policy Institute of Ohio, 2024, Ohio Department of Health, 2023) over the past few decades. The burden on patients, families, and their communities is immense, and the public health and economic costs have been overwhelming (Lou et al., 2021, Rockett et al., 2023,). Like other fields in medicine, some individuals from certain geographical areas (e.g. Appalachia, farmland, inner cities) and communities with difficult experiences (e.g. disadvantage via increased crime and pollution, decreased economic and educational opportunities, structural racism, and restricted health care access) have been disproportionately affected by overdose and suicide (Morales et al., 2020, Pamplin et al., 2023, White et al., 2024,). The COVID-19 pandemic has further exacerbated and highlighted these problems leading some investigators to identify avenues for improvement (Gupta et al., 2024, Parenteau et al., 2023, Radfar et al., 2021).

Unfortunately, efforts and strategies for prevention, treatment, and access to care have not yet resulted in a substantial population-level reduction of morbidity and mortality associated with these disorders (National Center for Health Statistics, 2024). This is perhaps not surprising, as we do not fully understand the root causes of psychiatric and substance use disorders, nor do we fully appreciate the benefits (and time sensitivity) of protective and buffering factors. Most studies in this area of research are conducted with individuals and in modality or diagnostic “silos”. For example, the focus of many investigations is on a single disorder, on a restricted number of risk or protective factors, on only a few modalities for inquiry (e.g. genetics, imaging), and often with small or modest sample sizes. Finally, the intergenerational transmission of risk for mental health conditions and adversity extends beyond genetics to environment, families, communities, and their interactions, which is also often siloed into separate disciplines and studies. In contrast, many factors are potentially modifiable (Huggard et al., 2023, Kamdar et al., 2023, Kirkbride et al., 2024), both from a risk and protective perspective.

Risks for, and protective factors buffering against, expression of mental health challenges are numerous and complex. Some factors related to overdose and suicidality are similar, while others (many not yet identified) may be either shared or unique. Notably, a substantial part of intergenerational transmission (of risk or resilience) relates to shared environment and parental behaviors, including abuse, neglect, and degree and type of supervision. For example, those who experience multiple adverse childhood experiences (ACEs) are up to 4-fold more likely to have substance use disorders, suicidality, and depression in large epidemiological studies (Anda et al., 2002, Choi et al., 2017, Copeland et al., 2023, Felitti et al., 1997,). In addition, deaths from suicide and drug overdose are often preceded by persistent psychological distress, which independently predicts both suicide (Copeland et al., 2020, Hockey et al., 2022, Stacy & Schulkin, 2022,) and drug overdose death (Betts, et al., 2015, Hockey et al., 2022, Shanahan et al., 2021) in) inin large prospective studies.

Many people who we lose to suicide and overdose have never had an interaction with a behavioral health service or system and have no mental health diagnosis, but <u>have</u> had interactions with health care systems (Luoma et al., 2002, Stene-Larsen et al., 2019). As such, it is imperative that health care systems have better understanding of ways in which we can support such individuals in terms of risk and resiliency factors. Our understanding to date is simple and linear and we do not yet have datasets of sufficient granularity and size to integrate multiple pathways to both risk and resilience. There are many possibilities and modalities to investigate.To attain a comprehensive and mechanistic understanding of the intricate pathways of risk and protective factors (and their interactions), a new strategy is necessary. Such a strategy necessitates larger samples, multimodal measurements, and embedded sampling (in families, communities, geographical contexts, etc). It also requires a longitudinal, temporal framework of study - how development, nurturing, and support are time-sensitive, unfolding, and interactive processes. Such comprehensive strategies hold great promise for transforming the way we conceptualize psychiatric and substance use disorders, and for how we can work to reduce suicides and overdoses. Such studies could also provide the necessary foundational framework to develop new strategies for identification, prevention, treatment, and monitoring that could lower risk and strengthen resilience at the individual, family, and community level.

Multimodal, longitudinal approaches have transformed other fields in medicine, where identification of risk factors at the individual level are leveraged to improve disease prevention at a population level, to develop individual level disease risk calculators, and to implement precision treatments tailored to an individual’s specific risk factors. Perhaps the most prominent example is the Framingham Heart Study (Andersson et al., 2019, Dawber et al., 1959). In 1948, this long-term, multigenerational cohort study recruited approximately 5,000 participants from a small community in Framingham, Massachusetts, with the objective to identify common factors that contribute to cardiovascular disease. The study has been running consecutively for 75 years, and knowledge has accrued incrementally. Specifically, this work (and subsequent work sparked by this study) has resulted in the identification of modifiable cardiovascular disease risk factors such as smoking, obesity, high blood pressure, high cholesterol, and physical inactivity. Notably, when the study began, some of these factors were considered static (not modifiable) and many of the outcomes were considered predestined - outside of the individual’s control. Framingham has resulted in thethethe development of individual-level risk calculators, numerous new prevention strategies, personalized treatment approaches, evolving understanding of what is modifiable, and new, sensitive disease detection and monitoring tools. These prior works about which protective factors are genetic and what factors are developed over time have illustrated just how much can be modifiable in the context of protection, or resilience.

Brain-based risk (and a few resilience) studies have begun to expand into the hundreds and thousands of participants. These larger studies have emerged in reaction to large scale studies of genetic risks (e.g. Mullins et al., 2022), which have identified hundreds of single nucleotide polymorphisms (SNPs) each carrying a small amount of risk for psychiatric illnesses, and many of these are from shared common genes. Large-scale deep-phenotyping studies aimed to understand risk for psychiatric and substance use disorders across multiple levels of analyses have also emerged. These studies are more expensive, and more comprehensive, yet almost all have held the individual person as the primary unit of study.A few prominent examples of longitudinal, imaging focused deep phenotyping studies include the Adolescent Brain Cognitive Development (ABCD) study (Bjork et al., 2017, Volkow et al., 2018), the Advancing Understanding of Recovery after Trauma (AURORA) study (McLean et al., 2020), the Philadelphia Neurodevelopmental Cohort (PNC) (Calkins et al., 2015, Satterthwaite et al., 2016), the Tulsa 1000 study (Victor et al., 2018), and the Texas Resilience Against Depression (T-RAD) (Trivedi et al., 2020) studies. While beyond the scope of this protocol paper, there are innovative design choices in these studies that informed our design of the State of Ohio Adversity and Resilience (SOAR) study. Interestingly, the T-RAD studies are one of a handful that have stated an explicit intent to study resilience factors. More recently, a consortium level approach called FAMILY (Karguth et al., 2024, van Houtum et al., 2024) builds upon workgroups and data fusion steps to understand intergenerational transmission of mental health challenges, predominantly from a genetic and neuroimaging perspective. . Most studies have held fast to the idea that the individual is the sole source of study at the level of brain and behavior, and have underweighted the importance of family, social, and environmental factors. Given the prominence of early life and family factors on risk and resilience, several (and larger) family-based studies will be extremely important to understand processes of intergenerational transmission and prevention. A direct intentional focus on aspects of resilience will also be critical.

There has been increasing interest in how individuals, families, and communities cultivate and develop resilience (Ayed et al., 2019, Ioannidis, et al, 2020, Métais et al., 2022, Schäfer et al., 2024, Windle et al., 2011,). Resilience is not the absence of risk, or even the absence of mental health challenges. Instead, resilience is the capacity to have positive and meaningful lives in the face or aftermath of exposure to adversity. Traditionally, resilience has been considered a developmental outcome of genetic and gene by environment process, whereby protective and buffering skills develop (*foundational* resilience) that protect against experienced risks. Additional conceptualizations of resilience have also emerged. *Emergent resilience* refers to a dynamic, adaptive process of “negotiating, managing, and adapting to significant sources of stress or trauma” facilitated by an interaction of internal cognitive-emotional capacities with social and environmental supports, accommodations, experiences and resources (Bessette et al., 2018, Windle, 2011,). For example, positive childhood experiences (PCEs), including positive interpersonal experiences, connectedness and sense of belonging with family, friends, and adults in social institutions and the community, that mitigate effects of adversity on mental health (Bethell et al., 2019) have been identified y. Education, economic resources and access to care, social capital, and faith and cultural factors also appear to be protective (Ayed et al., 2020, Flaskerud, 2022, Fuller et al., 2020,Windle, 2011). Excitingly, many potential emergent resilience factors appear to be modifiable with education, scaffolding, and/or support. S*ocial resource resilience* refers to the buffering and supportive factorsfactors and effectors thatthatresidingthat reside outside the individual. These supports are in systems, communities, and families.

The State of Ohio Adversity and Resilience (SOAR) study is intended to identify, understand, and exploit pathways to the development of all three aspects of resilience in a state-wide, population-based, family-linked multi-disciplinary, multi-modal longitudinal study of Ohioans. Ohio is considered a microcosm of the USA in many aspects (geography [farmland, cities, mountains], employment, education, race and ethnicity, religion, and variability in population density). As such, the state is ideally suited for conducting a broad generalizable research study of pathways to resilience. We aim to follow the SOAR cohort in regular two-year intervals over the next decade (and hopefully longer), similar to prior large-scale efforts in other disciplines, and will extend representative findings to an entire state (Ohio). The overarching goal of our study is to understand root causes and brain mechanisms of mental illness and substance use disorders, and to better understand intergenerational transmission of risk and resilience. To complete this work, we continue to build a statewide coalition of academic institutions, medical centers, hospitals, and community stakeholders. We will investigate social, environmental, psychological, and biological factors to map modifiable and non-modifiable risk and resilience signatures across multiple levels of analysis. The results of the SOAR study will create a novel foundational framework where prevention, treatments, and disease monitoring strategies will be developed based on multilevel risk and resilience factors. It aims to also provide participants, families, communities and lawmakers with actionable information to promote mental health and well-being. If successful, this comprehensive approach will allow us to “bend the curve” of morbidity and mortality related to psychiatric and substance use disorders.

SOAR also includes conceptual, pragmatic, and technical innovations. The study is designed to integrate with the ABCD study (Volkow et al., 2018) in technical measurements for some questionnaires, imaging techniques, and neuropsychological testing. The study also includes key innovations in methodologies for white matter and gray matter microstructural imaging (e.g., neurite orientation dispersion and density imaging, NODDI, Kraguljac et al., 2022), tests of inhibitory control and set shifting (Parametric Go/No-go and Balloon Analogue Risk Task, Lejuez et al., 2002), use of EEG, integration of multigenerational familial perspective, focus on aspects of resilience, and blended sampling strategies that include multiple geographic and population density frameworks at the state level.

Our first hypothesis is that we will be able to use predictive analytics to develop risk calculators / indices and resilience profiles at the level of groups, including key demographic, resource, and geographical factors. These calculators and profiles will include multimodal data from brain, behavioral, social, and individual measurements. Our second hypothesis is that these calculators and profiles, respectively, will include unique adjustments and perhaps even different features based upon a host of potential covariates, as noted above (e.g., sex, age, race, education, population density, economic opportunities). SOAR is designed as a data driven, exploratory, longitudinal study that can enable us to exploit and apply knowledge in real time to better the lives of Ohioans and to extend what we learn nationally and internationally.

## METHODS and ANALYSIS

### Overview of SOAR Design of Wellness Discovery and Brain Health Studies

The SOAR study consists of two principal investigations; targeted samples for the SOAR Wellness Discovery Survey of 15,000 Ohioans and the SOAR Brain Health Study of 3,600 Ohioans embedded in 1,200 families. After baseline assessments (detailed below), we aim to reassess the cohort at two-year intervals for generations to come.

The SOAR studies are designed to be two separate (and bidirectionally informative) investigations, assessing overlapping domains, with different sample sizes, sampling frameworks, and using different modalities. The SOAR **Wellness Discovery** study is designed to be broad-based screening of drivers of risk and protective factors as longitudinal, epidemiological sampling at the individual level. The SOAR **Brain Health** study is designed to have overlap in domains, questionnaires and longitudinal design with the Wellness Discovery Survey. It also expands to additional modalities of inquiry, referred to as deep phenotyping. We employ state of science approaches for multimodal measurement of brain structure and function, biomarkers, and psychological functioning, within a family-based approach (Figure 1 illustrates the sample frame and areas of study for the two studies over time).

**Figure 1.**
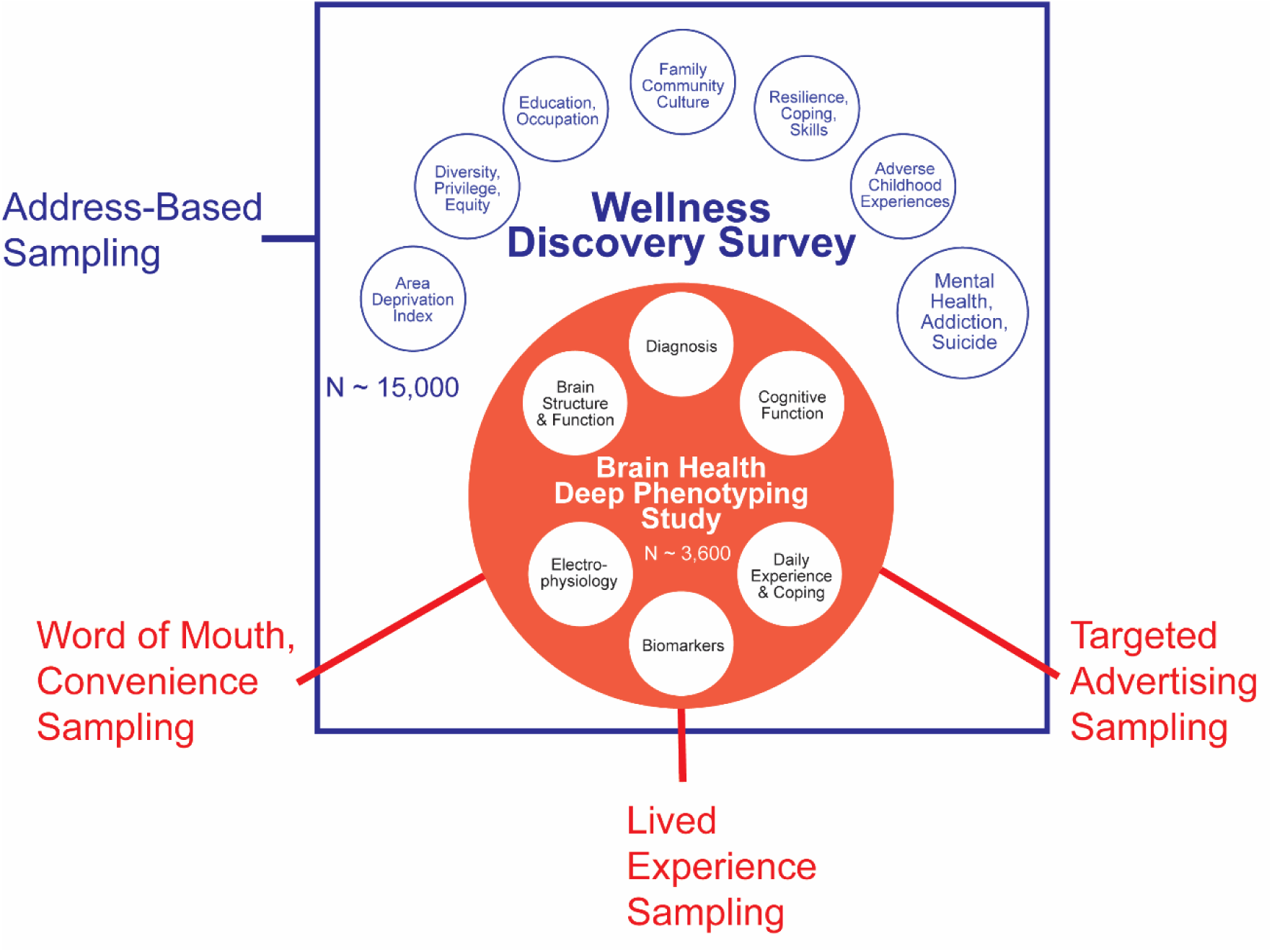
Illustration of how the SOAR Study includes an address-based sampling (ABS) sample representative of state of Ohio with broad light phenotyping Wellness Discovery Survey, and a Deep phenotyping, family-based Brain Health Study, which includes the Wellness Discovery Survey items plus additional questionnaires and multimodal biological, psychological and social measures.

The SOAR Wellness Discovery (WD) (WD)study includes address-based sampling (ABS) making use of a database listing (1) all people ages 18 and older from (2) all household addresses receiving mail in Ohio. Sampling zones are divided by geography, rurality, and demographics (Figure 2a) to enable weighted analyses that represent the entire state and can be extrapolated to regional, state and national subgroups.

**Figure 2.**
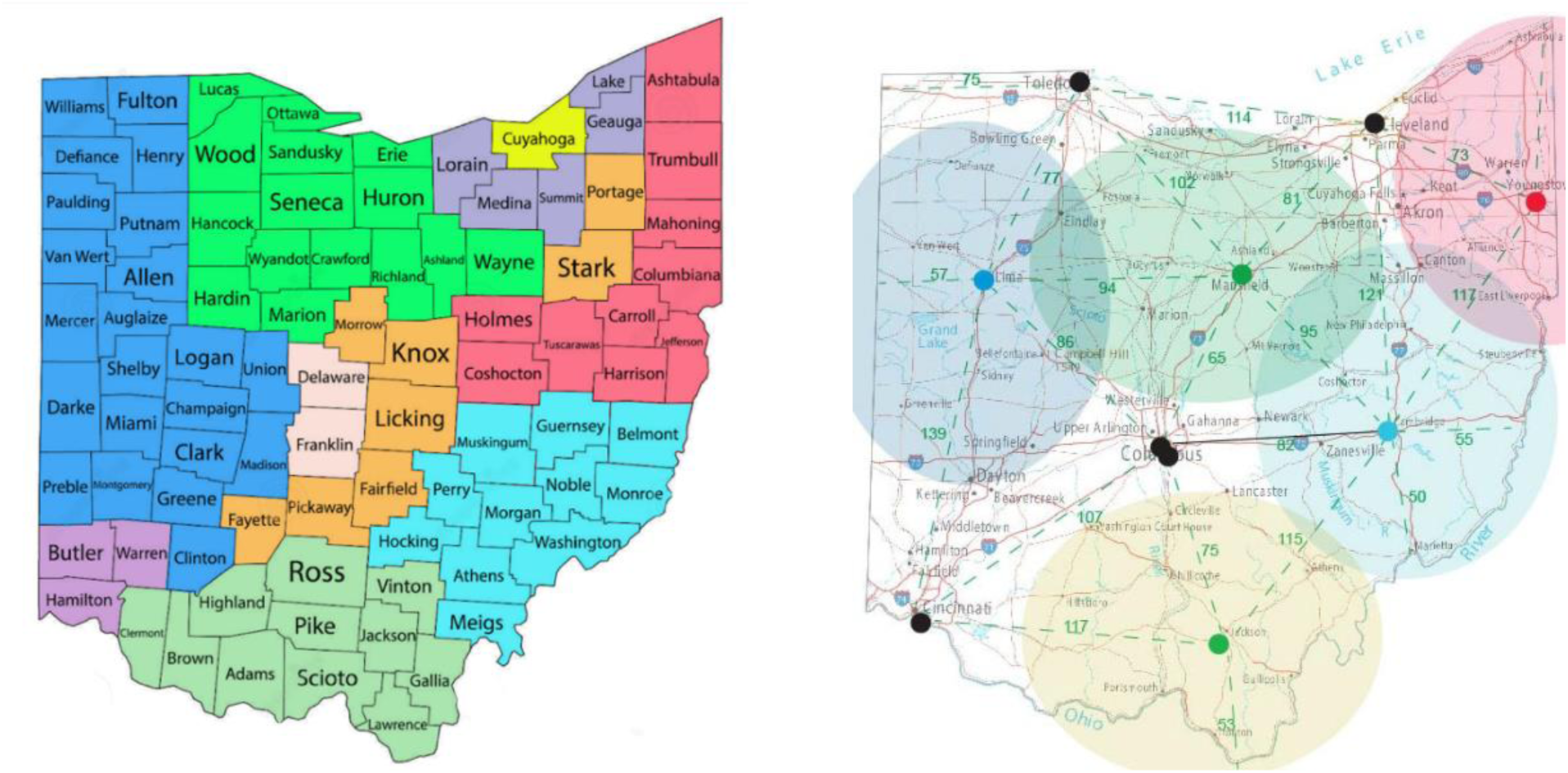
Panel A. Illustration of the sampling regions for WD survey, subdivided by urban – rural, geographic region, race and ethnicity. Urban counties are further subdivided to optimize sampling strategy (not illustrated here) Figure 2, Panel B. Illustration of the BH targeted mobile sampling sites (colored dots) and sampling zones. Green dashed lines and numbers indicate google mileage distances between locations. Black dots indicate stationary BH sites in medical centers. Mobile sampling sites vary pending contractual details.

The SOAR Brain Health (BH) study uses multimodal, deep phenotyping to carefully characterize up to 1,200 families and 3,600 individuals aged 12-72. The BH cohort will be comprised of participants from the SOAR Wellness Survey (address-based sampled). The sampling strategy also includes (1) convenience (2) word of mouth, and (3) targeted recruitment of individuals with psychiatric or substance use disorders, suicidality, or overdose (lived experience). These four strategies include concurrent co-recruitment of family members. The multifaceted strategy enables us to compare and contrast with other types of longitudinal studies that use one, but not all of these sampling strategies.

The strategy to include those with lived experience enhances the relevance of the work. In contrast, the address-based sampling, convenience, and word of mouth sampling align with other research frameworks where individuals may tend to be more high functioning. SOAR’s unique BH sampling strategy includes use of both stationary existing research MRI capable sites AND use of a mobile assessment strategy partnering with regional hospitals (Figure 2B). The mobile research capable MRI scanner spends 2-4 months at each recruitment site. This innovative strategy enables any Ohio family to participate in the BH study by traveling less than 80 miles, enhancing access and representativeness of the sample (particularly for rural participation).

The timeline of the SOAR Studies data collection Waves started in 2023, with the start of the BH Wave 1.0 in November 2023 and the WD Wave 1.0 in January 2024. We anticipate Wave 2.0 to initiate in the fourth quarter of 2025, and for data collection of Wave 2.0 to continue over two years. Each subsequent SOAR Wave of data collection is planned to be conducted over a 2-year (∼24 month) period.

**Figure 3.**
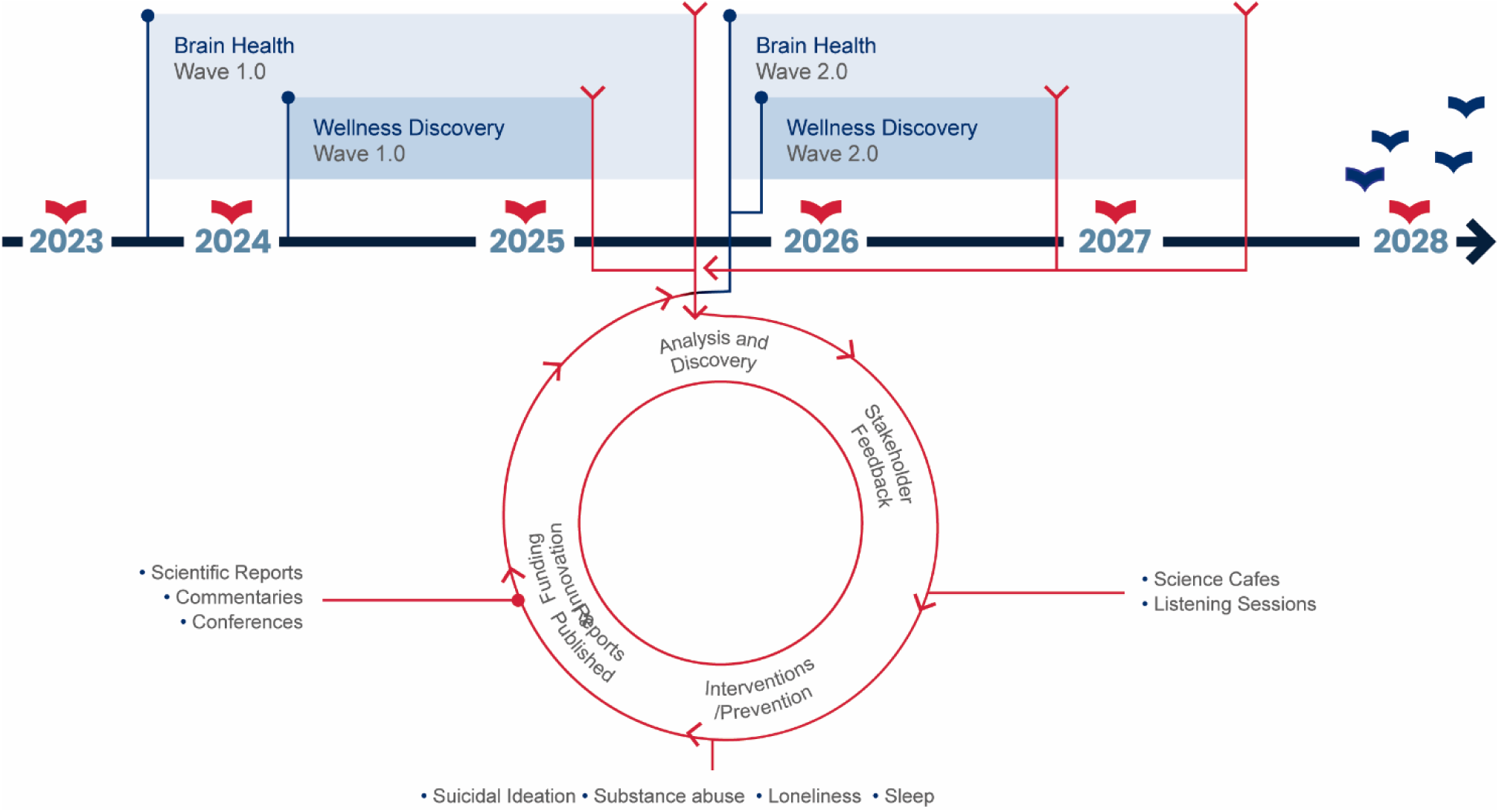
Anticipated timeline of the SOAR Studies – Wellness Discovery and Brain Health Waves 1.0, and Wave 2.0. Waves 3.0 and beyond not shown and all are contingent on fundingI.

Together, this multifaceted approach (Figure 1) gives us the necessary breadth and depth needed to comprehensively capture modifiable and stable aspects of risk and resilience factors in a longitudinal, interactive framework.

### The SOAR Study and SOAR Network of Academic and State Partners

The SOAR Study is directly commissioned by the Ohio Department of Mental Health and Addiction Services (OhioMHAS) through a State of Ohio legislative bill. Administratively, the OhioMHAS has also spearheaded a parallel SOAR Network, which is intended to elicit immediate action interventions with community boards and behavioral health providers and organizations directly. The network’s aims are “to innovate through taking results of applied research and disseminating them more broadly so that more people understand and are prepared to implement strategies that help prevent mental illness and substance use disorders, more effectively treat these illnesses, and support long-term recovery for those experiencing the conditions”

The SOAR Study is anchored at The Ohio State University (OSU) Wexner Medical Center and conducted in partnership with academic, community, and government partners across Ohio. Academic partners include: Bowling Green State University, Central State University, Kent State University, Nationwide Children’s Hospital, Northeast Ohio Medical University, Ohio University, University of Cincinnati, Cincinnati Children’s Hospital Medical Center, University Hospitals/Case Western Reserve University, University of Toledo, and Wright State University. Partners also include behavioral health providers and community hospitals (e.g. Avita Health Systems in Ontario, OH and Holzer Health in Jackon, OH), community organizations (active and ongoing discussions and coordination with One Ohio Recovery Foundation, Ohio Suicide Prevention Foundation, and related community organizations), and local Alcohol, Drug and Mental Health (ADAMH) boards. Partners interact, guide, and lead SOAR activities in workgroups, events, community engagement, dissemination of knowledge gained, and other activities relevant to the mission of the SOAR studies.

### SOAR Organizational Structure

We have assembled Cores that have designed and implemented the SOAR assessment instruments and procedures for data acquisition and have facilitated the emergent academic collaboration for both data acquisition and scientific discovery, a Safety and Ethics Committee, and Workgroups for Data Access and Community Engagement.

#### Administrative Core

The administrative core includes the steering committee for the study, and other necessary administrative support. The Steering committee directs the collection, administration, and reporting of the study data. The steering committee directs the workgroups and committees and other Cores to meet SOAR study objectives.

#### Wellness Discovery Survey Core

The Wellness Discovery Survey is fielded by the Center for Human Resource Research (CHRR), a nationally recognized survey research center housed at OSU, with a subcontracted mailing organization and subcontracted call center in Chillicothe, Ohio.

#### Neurophenotyping Core

The neurophenotyping core includes key area experts who have designed and codified the manual of procedures for data acquisition using our multimodality strategy. These include subgroups that have optimized procedures and data acquisition, training, and quality control procedures for stationary 3T and mobile 1.5T MRI scanners,, EEG data collection, ecological momentary assessment (EMA), neuropsychological testing, diagnostic interviewing, and questionnaires of demographics, familial and social factors, resilience factors, adversity factors, substance use and mental health conditions.

#### Biomarkers Core

This core is dedicated to safe biospecimen collection, processing, and secure storage of biospecimens for future planned biomarker assays and inclusion of these data into the SOAR analyses.

#### Data Management Core

The data management core has designed the data management plan. The core creates and maintains an Integrated Data Storage System that contains a linked dataset from the SOAR studies. The core manages the unification of both data and metadata into a single repository for engagement, linkage, migration pathways to a single computer platform, and long-term storage. Metadata management on behalf of the study will be done via an Honest Broker system. Data access will be provided following approval by the Data AccessAccess, Publication and Presentation Workgroup. Requests for access to Personally Identifiable Information (PII, e.g. zip code, address, phone number) from partner sites will require additional Data Use Agreements and IRB approval. Documentation and management of access are actively monitored, with access reviews occuringoccuring on a quarterly basis.

#### Biostatistics Core

The Biostatistics core has responsibility for guiding data cleaning and data processing steps, honing hypotheses, designing data analytic techniques, and implementing data analysis steps.

### Committees

#### Safety and Ethics Committee

We expect to have anticipated (and unanticipated) adverse events, as the samples include base rates of such events. The committee evaluates standard operating procedures for management of adverse events and review cumulative study data to evaluate safety, study conduct, and ethical principles of the study. It provides recommendations to the PI and administrative core leaders. The Committee will meet twice a year and consist of three members with a current appointment at any of the participating institutions, but without any other role(s) on the study. In addition, *ad hoc* meetings can be convened at the PI’s request - a review in case of a severe adverse event has occurred. Individual terms are 3 years, and; these are staggered to allow for continuity.

### Workgroups

#### Data Access, Publication and Presentation Workgroup

This workgroup will oversee and monitor access to SOAR data and to facilitate the production of high-quality scientific publications and presentations on behalf of SOAR according to the Publication and Presentation Policy. The workgroup is responsible for overseeing, reviewing and tracking of SOAR data requests, study manuscripts from inception to publication, and study presentations at scientific conferences. The workgroup will ensure that the necessary regulatory approvals are in place but will not provide support for data analytic or dissemination activities or logistics and administrative support to writing teams. Lead authors assume responsibility for these activities. Each academic center will have a representative on the workgroup whichwhichwhichwith 2-year terms that arestaggered for continuity, and the workgroup will have. TTTerms. The workgroup will have monthly meetings.

<u>Community Engagement Workgroup</u> identifies and meets with local and regional community organizations, stakeholders and leaders across the state of Ohio. The engagement activities center around building awareness of the SOAR study, listening and learning from communities about their health priorities and local context, and building trusted regional networks of partners in the state of Ohio (Aguilar-Gaxiola et al., 2022). In studying the drivers of health and building a network of partners, SOAR seeks to build bi-directional accessible communication and dissemination of findings with all community stakeholdersand. Ohio has mental health services administered through the Ohio Mental Health and Addictions Services (Ohio MHAS), and this includes 50 regional addiction, drug and mental health boards (ADAMH boards) divided at county or multicounty levels..

### SOAR Wellness Discovery (WD) Study Protocol

#### Participants

A random address-based sample, representative of the State of Ohio, of 15,000 adult people who reside in Ohio, receive mail, and have the ability to complete the survey. (Figure 2A).

#### Recruitment

Multiple waves of mailing of 300,000 postcards (and subsequent targeted mailings by region, age, sex) are conducted. Targeted mailings include unique QR codes for the targeted individual within the household. A call center also works to contact individuals directly based upon the location and demographics (e.g., males and older individuals are more likely to respond to calls).

#### Informed consent

The IRB of record for the Wellness Discovery Survey is The Ohio State University IRB. Written informed consent is obtained virtually and recorded in the CHRR platform. After completion of the survey, participants are given the option to agree to be recontacted for the Brain Health Study. Consent also includes authorization to connect information to the SOAR Wellness Discovery survey (if completed) responses and to publicly available state databases (e.g. Vital Statistics data, Medicare claims data, data from the Ohio Longitudinal Data Archive) to enrich the dataset and provide complementary data on longitudinal outcomes.

#### Inclusion/ Exclusion criteria

All adults in Ohio are eligible using address–based sampling and capability to complete the questionnaires via online survey or telephone interview in English..

#### Assessments

The QR code provides access to the survey for online completion. Alternatively, participants can complete the survey via a phone line that will connect the participant to study staff who will assist in completion of the survey over the phone. The survey takes about 50 minutes to complete and includes questions related to demographics, life experiences, mental health challenges, education, occupational activities, economic outlook, resilience and coping skills, social functioning, and environments (see Figure 1, Panel A, and Table 1).

**Table 1.**
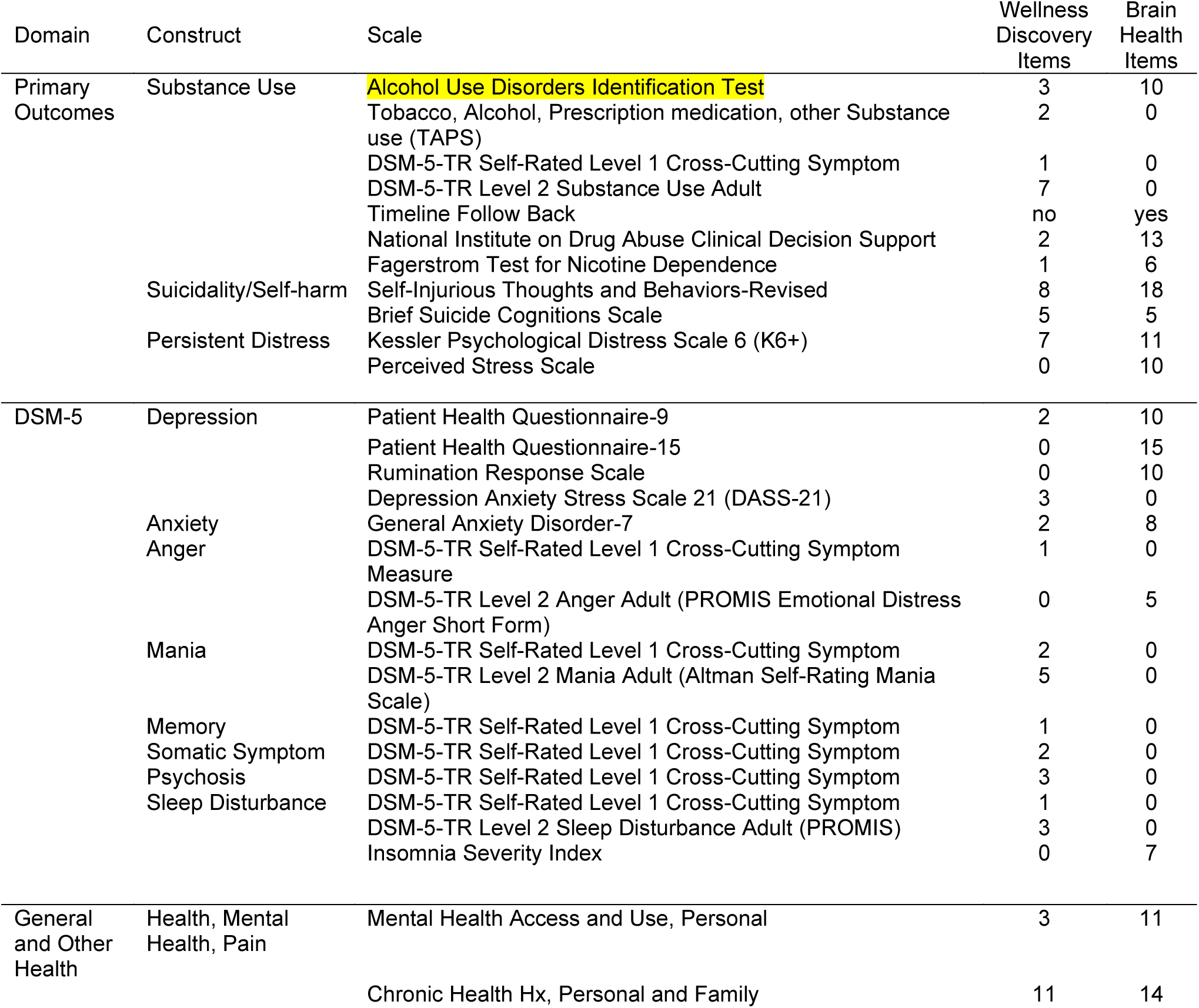
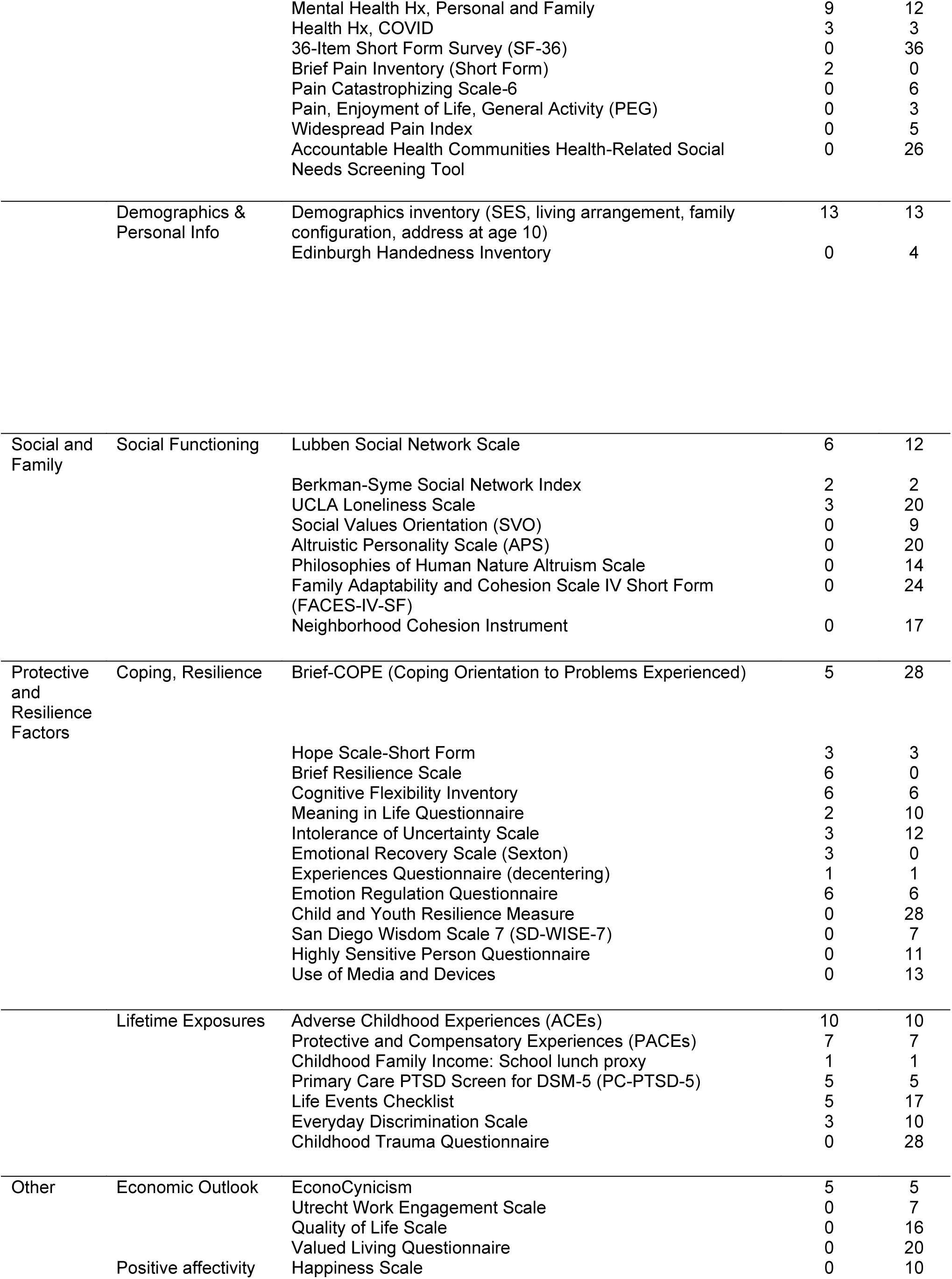
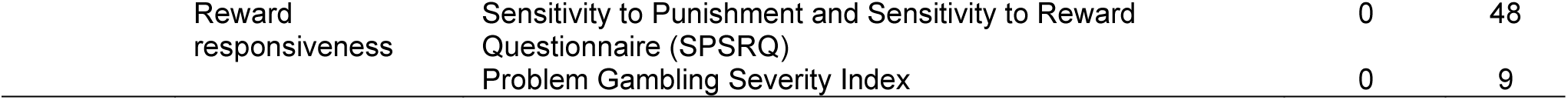
Assessment and Survey items in the Wellness Discovery Survey and Brain Health Studies. . Shown are the domains, constructs, and specific scales, and number of items in the Wellness Survey and Brain Health Study

#### Compensation

ParticipantsParticipants will be compensated for their time at a rate of approximately $1 per minute for completed questionnaires, up to $50, in the form of electronic or physical gift cards.

#### Data analysis plan for Wellness Discovery Survey

The biostatistics core, interested workgroups, and steering committee will work in unison to analyze the WD data. The size and address-based sample enable key analytic steps. First, weighting will be conducted based upon region (sampling cluster, see Figure 2a) and actual demographic data from the 2020 census. Second, data cleaning steps will be employed to identify and adjust for skewed or out of range values. Third, data aggregation steps will be developed to create adversity and resilience index(es) and other predictive analytics of development of substance use problems, suicidal thoughts and behaviors, and persistent psychological distress (our three primary outcome variables). Such indices will be evaluated from invariance across region, sex, race, religious, educational and economic background, as it is possible (and indeed likely), that tuning of such indexes is necessary to enhance precision in detection, programmatic targeting, and individualized outcomes. These large group indexes can then be used in subsequent analyses with the BH study (below).

### SOAR Brain Health (BH) Study Protocol

#### Participants

The Brain Health Study will recruit 1,200 families (∼3,600 individuals) aged 12-72.

The first adult member of a family who agrees to participate in the study is asked to invite 1-5 additional family members to participate (average 3 people per family; see Figure 1). The Brain Health cohort will include participants from the SOAR Wellness Discovery Survey, a convenience sample of randomly selected families, and a nested sample of families with members with a history of psychiatric or substance use disorders, suicidality, or unintentional drug overdose..

#### Recruitment

Individuals are recruited across the state of Ohio. Families are not required to be genetically related, but rather are “family units” as self-defined, and thus can include “blended” families and fictive kin/family of choice. Recruitment strategies include linked recruitment through the WD study (an interest question at the end of survey authorizing contact), direct connections through community organizations, and direct advertisement through distribution of flyers, posterboards and other informational materials, ads on Facebook, hosting of community engagement events, and participation at local events.

#### Informed consent

A single IRB was used, centralized at OSU. Consent is conducted either via virtual or in person visits (if information is collected in person, a local consent is also signed, e.g., at University of Cincinnati). Consent also includes authorization to connect information to responses provided in the SOAR WD study (if completed) and to publicly available state databases (e.g. Vital Statistics data, Medicare claims data, data from the Ohio Longitudinal Data Archive) to enrich the dataset and provide complementary data on longitudinal outcomes..

#### Inclusion/ Exclusion criteria

Individuals aged12 and above who reside in Ohio are eligible for study participation. Exclusions are procedure specific based upon safety (e.g., no MRI if pacemaker is present), and comprehension (understanding study procedures and ability to complete the protocol step).

#### Study sites

Data collection begins at 6 academic medical centers, Case Western Reserve University/University Hospitals, Nationwide Children’s Hospital, The Ohio State University Wexner Medical Center, the University of Toledo, the University of Cincinnati/Cincinnati Children’s Hospital Medical Center. These academic medical centers were selected based upon 3T MRI capability, geographic dispersion, catchment area, local infrastructure for MRI scanning and other deep phenotyping assessments, and expertise to complete study procedures.

In addition, data are also done using mobile sample strategies to minimize burden for participants residing >80 miles from a stationary site, which includes Ohio’s Appalachian region and Ohio’s farmland regions. We deploy a mobile laboratory with a 1.5 Tesla Siemens Mobile Avanto mobile MRI scanner hosted at distributed community hospital partner site for 2-4 months. Sites are selected based upon geographical location, interest in participation, capability to accommodate space, electrical, internet, and phlebotomy requirements for the mobile scanner, and availability of facilities that can be used to facilitate deep phenotyping data collection other than MRI imaging.

#### Assessments

Assessments are conducted in a modular fashion. Visit days, times, module number, and order are adjusted to accommodate participant schedules. Participants can opt out of individual modules by interest, comfort, safety, and availability. For example, if a participant cannot tolerate an MRI scan or is averse to blood draws, they can choose to still participate in the overall BH study, but not complete specific modules. To ensure participant comfort and avoid excessive fatigue, there is an option to split study modules up across visits. Some components can be completed entirely remotely (diagnostics, questions, neuropsychological testing, diagnostic interviews).

#### Diagnostic interview

We use the quick-SCID (First & Williams, 2020), a structured clinical interview for DSM-5, conducted by trained interviewers completed in person or virtually (Zoom, Teams). There is strategic priority ordering (minors, family members of minors, those with intermediate DSM-5 screener scores, those reporting current and prior exposure to potential trauma and abuse),),), and a random sampling strategy) until all individuals have completed the interviews.

#### Questionnaires

Participants complete a broader set of questions and questionnaires (that include the WD survey) in REDCap. Table 1 illustrates the comparative and integrative sampling strategies of the WD surveys and the BH surveys.

#### Anthropometrics

Height and weight are measured.

#### Neuropsychological Testing

Cognitive testing is completed to align with key elements of longitudinal adolescent brain cognitive development of youth (e.g., NIH Toolbox, working memory, set shifting, processing speed, working and verbal memory, https://abcdstudy.org/). The Flanker Inhibitory

Control and Attention test assesses inhibitory control and attention. The List Sorting Working Memory Test assesses working memory abilities, including ability to hold items in memory briefly and complete sorting operations on those items in working memory. Pattern Comparison ProcessingProcessing Speed isa used for assessing and responding based upon a sorting perception rule. The Picture Sequence Memory Test assesses memory for integrated auditory and visual order information and the Rey Verbal Learning Test assesses learning of serially presented words. The Oral Reading Recognition is an NIH task assessing reading decoding skills and crystallized abilities. Matrix Reasoning is used as an IQ estimate and Finger Oscillation is included for a processing speed attenuation component. Cash choice is a single item delayed discounting question. We also include high-fidelity tasks related to suicide and addiction (impulsivity and inhibition with Parametric Go/No-go and Balloon Analog Risk Tests completed through Pavlovia system on iPad tablets).).

#### Multimodal MRI

Magnetic Resonance Imaging is conducted using multiple platforms at five stationary sites at academic medical centers, and a mobile MRI scanner which is initially to be deployed sequentially at four sites across the state of Ohio. At OSU, scanning occurs at the Center for NeuroImaging, NeuroPhenotyping, NeuroComputation, and NeuroModulation (C4N), a new research-dedicated shared facility for human clinical and translational studies in the OSU Department of Psychiatry and Behavioral Health. Scan sequences are aligned with the ABCD longitudinal study.

Scanning is conducted using a Siemens MAGNETOM Cima.X 3T MRI scanner with Gemini Gradients 200* mT/m @ 200 T/m/s, which runs on syngo Expert-i XA60/XA61a software. Sequence packages include Neuro fMRI/DTI, Arterial Spin Labeling 3D, pCASL, spectroscopy and SVS spectral editing packages. At Case Western Reserve / University Hospitals scanning is completed at CWRU Center for Imaging Research (CCIR) using a Siemens 3T Vida MRI scanner. At the University of Cincinnati College of Medicine, scanning is completed at the Imaging Research Center at Cincinnati Children’s Hospital Medical Center Imaging Research Center using a Philips 3T 7700 MRI scanner. Scanning is completed at Nationwide Children’s Hospital Department of Radiology using a Siemens 3T Prisma MRI scanner, at the University of Toledo at the ECORE Imaging facility using a GE 3T SIGNA Premier MRI scanner. Scanning at the mobile scanner is using a 1.5 T Siemens Avanto MRI scanner. Each site performs multimodal MRI scanning for assessment of brain structure and function. **<u>T1- and T2-weighted Structural scans</u>** are conducted for high resolution structural brain imaging: T1w (1×1×1 mm^3^/vox resolution) and T2w (1×1×1 mm^3^/vox resolution) to allow for cortical and subcortical volumetric analyses, grey matter density (e.g. voxel-based morphometry, VBM) analyses, and cortical thickness analyses. **<u>Diffusion-weighted imaging (DWI) scans</u>** using neurite orientation dispersion and density imaging (NODDI, 2×2×2 mm^3^/vox resolution) is conducted for analyses of cellular tissue architecture and gray and white matter microstructural integrity and abnormalities (Kraguljac et al., 2022) (e.g. patterns of neurite density and fiber orientation and microglial density), which may provide metrics of neuroplasticity and neuroinflammation. A custom analysis pipeline designed for SOAR by NVK using the NODDI toolbox will generate imaging maps and calculate NODDI metrics of neurite density (ND), orientation dispersion (ODI), and extracellular free water (FW). **<u>Functional MRI scans</u>** are conducted for blood oxygen level dependent (BOLD) analyses of brain functional activity and functional connectivity and are built to harmonize with the ABCD templates across 3T platforms (Casey et al., 2018). Resolution of fMRI scans are 2.4×2.4×2.4 mm^3^/voxel resolution for the stationary sites and 3.5×3.5×3.5 mm^3^/vox resolution for the mobile. **<u>Resting state</u>** scans (eyes open, two scans ∼6 minutes each are performed to allow for resting state functional connectivity analyses (e.g. DeJoseph et al., 2022). **<u>The Rumination Induction</u>** task (RIT) is utilized to assess neural correlates of negative rumination as well as positively valanced “savoring”. The RIT is a reliable and sensitive block design task that includes 4 self-relevant rumination prompts and 4 distraction-oriented imagery neutral imagery prompts (Cooney et al., 2010, Schreiner et al., 2023). It is modified to also include two savoring prompts/sequences. Conditions used for analysis are in 30 second blocks, including memory recall reminders, rumination experience and processing, and distraction conditions. Jittered rest blocks of 8-12 seconds are interspersed to add temporal power, and rumination and sadness / happiness queries are included after each rumination, savoring, or distraction block. The RIT fMRI consists of two scans at ∼6 minutes 30 seconds and the sequence parameters are aligned to the ABCD template as published in our prior work (Burkhouse et al., 2016). The total amount of scan time is ∼52 minutes.

All MRI data collected by SOAR will be shared and managed through an integrated, standardized data management system designed by the SOAR Data Management Core (led by authors TRH and NVK) and securely stored at the Ohio Supercomputing Center. Mode-specific quality control / assurance, pre-processing, and feature generation and analyses will be performed using custom standardized analytic pipelines designed by SOAR Neuroimaging Core (led by NVK).). SOAR will utilize state of the art harmonization tools to align stationary site data. Mobile 1.5 T data will be harmonized to 3T data as possible, but for some analyses, 1.5 and 3T data analyses will be conducted separately. Details of the standardized SOAR data management system and multi-modal MRI data QC, pre-processing, and feature generation / analytic pipelines will be described at length in a separate publication.

#### Electroencephalography (EEG)

We will record continuous noninvasive EEG using a 32 channel ActiveTwo BioSemi (BioSemi, Amsterdam, Netherlands) EEG recording system. Recordings will be collected during rest and up to four tasks (Table 2). (1) **Error Monitoring (Flanker) Task.** The modified arrowhead version of the original flanker task (<u>Eriksen and Eriksen, 1974</u>) is used to measure neural activity to error and correct responses (i.e., the ERN and correct response negativity [CRN], respectively). EEG to assess the Error (Participants see string of arrows on the computer screen and are asked to indicate which direction the center arrow is pointing using a computer mouse (i.e., left or right). Participants are told to make a rapid and accurate decision regarding the center arrow and respond quickly and accurately. (2) **Response to Acute and Sustained Threat. TThreatThreat.** This task, adapted from Grillon and colleagues’ (Schmitz & Grillon, 2012) No-, Predictable-, Unpredictable –Threat (NPU) paradigm captures reactivity to the anticipation of predictable and unpredictable threats. Startle eyeblink potentiation electromyography [EMG]) is recorded throughout the task as an index of aversive responding via electrodes placed over the orbicularis oculi muscle. In this version of the task, the ‘threat’ is an 85 dB aversive female scream noise. There are three within-subjects task conditions. In the “No-threat” (N) condition, no screamsare delivered, in the “Predictable threat” (P) condition, screams are delivered when a numeric countdown reaches “1.” In the “Unpredictable threat” (U) condition, threats are delivered at random. (3) **Doors Reward Task.** This task measures reward responsiveness via the reward positivity (RewP; also referred to as feedback negativity [FN]) of neural responses to monetary gain and loss, respectively. Participants are presented with two “Doors” to choose from, which gives chances for monetary gain or loss.Participants are told they can win up to $5 in the task; all participants are paid $5 embedded within the EEG payment.

**Table 2.** EEG Tasks and Potentials. Shown are the psychological domain, the event related potential (ERN) measured, and the behavioral task of each EEG assessment.

| Table 2. EEG Tasks and Potentials |  |  |
| --- | --- | --- |
| Psychological Domain | Event Related Potential or measure | EEG task |
| Error processing | Error-related negativity (ERN,) | Flanker task |
| Threat processing | Fear- and anxiety-potentiated startle | No threat, Predictable threat, Unpredictable threat (NPU) |
| Reward Processing | Reward Positivity (RewP) | Monetary Choice ("Doors") |
| Emotional expression and emotional regulation | Late Positive Potential (LPP) | Emotional Regulation Task |

**(4) Emotion Regulation** This ERP task measures late positive potential (LPP) (Fitzgerald et al., 2017) following viewing developmentally appropriate emotionally-valenced pictures (40 neutral, 40 unpleasant, and 40 pleasant) from validated published stimulus sets. With instruction prior to each block, the protocol involves five tasks: “Look”-Neutral, “Look”-Negative, “Look”-Positive, “Decrease”-Negative, and “Increase”-Positive, block order pseudorandomized within and across subjects. In the ‘Look’ condition, the subjects passively view neutral, positive, or negative pictures without any explicit attempt to regulate the absence of affect. During the ‘Decrease’ condition, participants are instructed to voluntarily decrease the intensity of their negative affect in response to negative images by using cognitive reappraisal. During the ‘Increase” condition, participants are instructed to voluntarily increase the intensity of their positive affect in response to positive images by using savoring/positive regulation strategies.

#### Biospecimen collection

Biospecimens will be collected and archived at -80⁰ C for future downstream analyses of whole-blood leukocyte DNA, RNA, and plasma biomarkers of stress response, inflammation, and metabolic function. Standard phlebotomy procedures will be employed to safely obtain venous blood specimens, a total of 45 ml into EDTA vacutainers and a TEMPUS™ tube (for whole blood leukocyte RNA). A whole blood specimen (4 mL) is frozen for DNA, the remaining blood is centrifuged at 2000 RCF to separate blood components, and multiple aliquots of plasma and “buffy coat” (white blood cells / whole blood leukocytes) are frozen and securely archived at -80⁰ C. We will also perform point-of-contact assays for Hemoglobin A1c and lipid profiles (HDL, LDL, total cholesterol, triglycerides, blood glucose) using benchtop analyzers (Abbott Labs, Infinion and Cholestech LDX) from collected venous blood or fingerpricka.

#### Ecological Momentary Assessments (EMA) and digital phenotyping

A subset of participantsparticipantsparticipants will complete 28 days of ecological momentary assessment (EMA) delivered using the HIPAA compliant, cross-platform (i.e., iOS and Android) MetricWire application. Participants receive 5 prompts / day for total 140 EMAs per participant. A static morning survey will be administered and then EMAs will be sent at pseudorandom times 4x/day, at least 3 hours apart, based on participant’s sleep/wake times. Items will assess SOAR primary outcomes (distress, suicide risk, substance use) as well as risk and resilience factors. EMA surveys will include an additional rotating battery to randomly select items from a broader question bank of risk and resilience factors to capture sufficient frequency (∼3x/day per item), while reducing participant burden. Adolescent and adult batteries will include largely overlapping items with some adolescent specific items (Table 3). An integrated safety protocol will direct participants to contact emergency resources if they endorse severe suicide intent or recent suicidal behavior during EMA. A list of mental health resources is available in the application at all times.

**Table 3.**
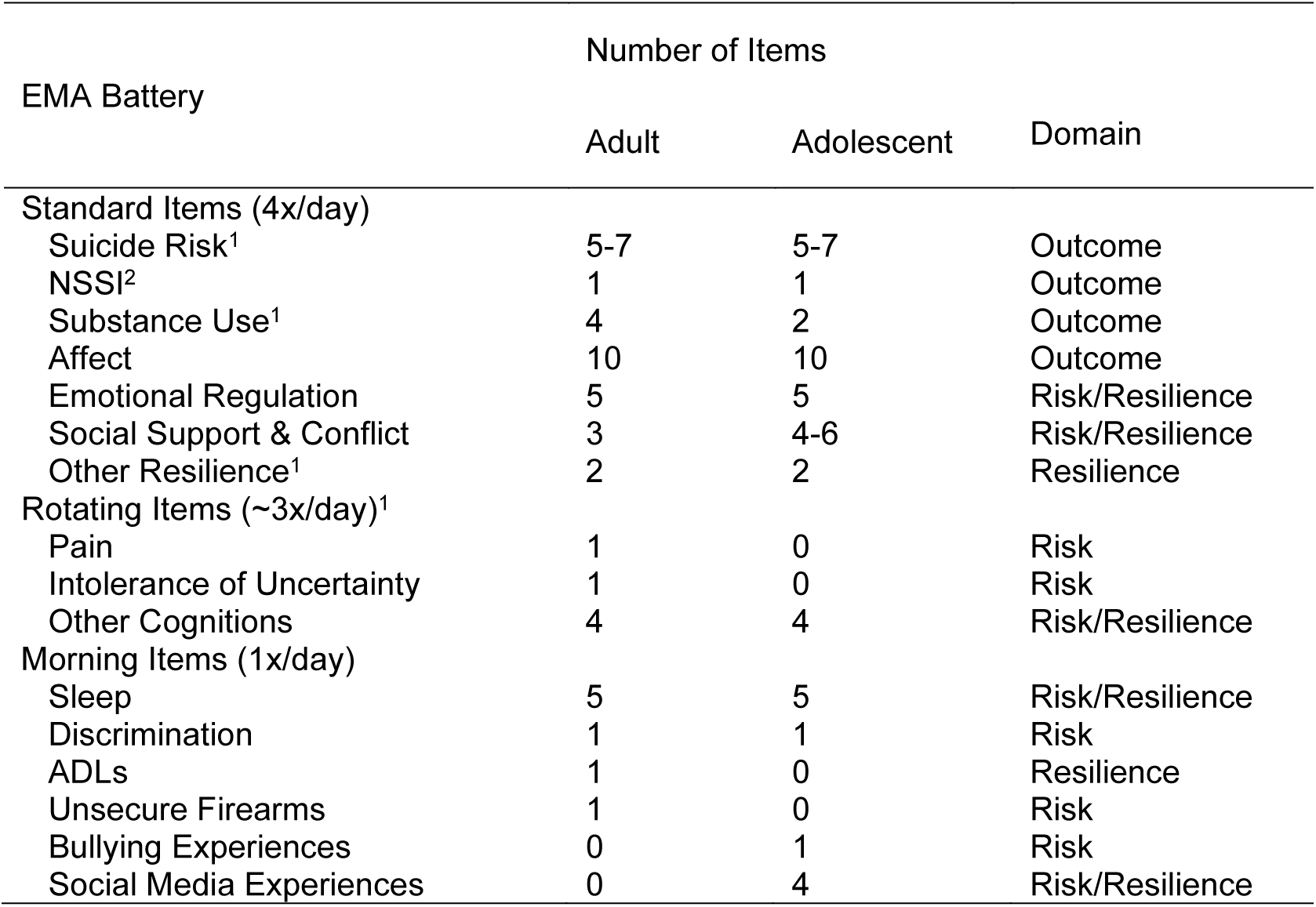
Ecological Momentary Assessment Data Collection module. ^1^Item also delivered during morning battery; ^2^Item delivered during standard and morning battery for adolescents, but during morning only for adults. Abbreviations: NSSI, Non-suicidal self-injury; ADL, Activities of Daily Living

#### *Passive* Smartphone Sensing *sensing*

WeWeWe will also passively collect mobile sensor data for digital phenotyping. Derived data include ambient light and noise, tracking nearby Bluetooth devices, call/text logs, app usage, pedometer (i.e., steps), and physical activity (i.e., stationary, walking, in transit) tracking based on accelerometry. All EMA and sensor data collected arede-identified. Participants may decline permission to collect each type of sensor data.

#### Data Analysis of the Brain Health (BH) Study

Core analyses are presented briefly here. Similar to those used in the oneswhat is the Wellness Discovery (WD) analyses, multimodal aggregate adversity indexes will be created by location, by family, and by individual using demographic, economic, personal measures and related social determinants of health (SDoH). Aggregate resilience and coping indexes will be created based upon location, by family and social network and by individuals (multimodal data). Data reduction steps for high and low dimensional data are often demanding, multistep processes (Calkins et al., 2015). Power analyses in longitudinal, multimodal data sampling studies are difficult to compute. Within subject, mixed level models are incredibly powerful in this context and accommodate significant data loss while retaining generalizability.

BH and WD measures will be included as exploratory and potentially modifiable predictors of cross-sectional and planned future longitudinal outcomes. For example, previous work by our teams has used EEG and MRI data to predict longitudinal outcomes in trauma, addiction, psychosis, depression, and treatment contexts. Longitudinal analyses address predictors of mental health outcomes within persistent distress domains, including anxiety, depression, trauma, psychosis, and substance use. We will have data related to 30-day history of substance use and 30-day to lifetime history of suicidal thoughts and behaviors. Suicide and overdose losses in the state of Ohio (15.0 per 100,000 (Health Policy Institute of Ohio, 2024) and 39 per 100,000 (Ohio Department of Health, 2023), respectively) are unlikely to be of sufficient number for logistic regression longitudinal models. In contrast, there were >30,000 (∼272 per 100,000) emergency department (ED) visits in Ohio related to drug overdose crises in 2023 (Ohio Department of Health, 2023) and extrapolating national trends suggests another ∼40 per 100,000 ED visits for crises related to suicidal ideation or behaviors, suggesting such crises may be able to be statistically analyzed. The family design of the BH study, including genetically related parents and offspring (as well as genetically unrelated caregivers and children) will also allow for future exploratory studies of intergenerational transmission of risk and resilience factors, including approaches such as family-based association testing.

Alignment and potential for harmonization with ABCD data provides another powerful potential avenue for our deep phenotyping models to predict outcomes and for these to be replicated in ABCD data, even if there is lighter clinical phenotyping and familial context measures from the ABCD study. There is overlap in neuropsychological measures, MRI measures, biospecimens, and questionnaires. SOAR data and analyses can enhance what is learned in the ABCD study, by including intergenerational, familial, and social contextual measurements.

## Data Availability

This is a protocol paper. All data produced in the present description of the protocol are contained in the manuscript.

## ETHICS AND DISSEMINATION

Ethical and safety considerations, as described above, will be managed by our Administrative Core, our Safety and Ethics Committee, and The Ohio State University Institutional Review Board.

Our dissemination plan will include scientific publications and white papers reporting data and results of analyses to the scientific and policy communities. We also plan for high-level reporting of data and results to stakeholders and the public in accessible manners, including reports and report summaries, and web-based reporting at appropriate reading levels.

All data collected as part of the SOAR studies will be processed and curated by our Data Management Core according to our comprehensive data management plan. High density data modality sources (e.g. structural MRI, NODDI, functional MRI, and EEG recordings) will have quality control measures applied and be processed through standardized analytic pipelines designed and implemented by our Data Management Core. A validated dataset of merged multi-modal data (self-report, interview, and testing responses, and data elements and features extracted or derived from MRI and EEG recordings, and biomarker assays) will be archived at The Ohio Supercomputing Center and the Ohio MHAS platform.

## DATA ACCESS STATEMENT

Technical reports and appendices, statistical code, and datasets will be made available to qualified researcher from the State of Ohio Mental Health and Addiction Services and The Ohio State University.

